# Pharmacological interventions for COVID-19: Protocol for a Rapid Living Systematic Review with network meta-analysis

**DOI:** 10.1101/2020.05.02.20088823

**Authors:** Ana Carolina Pereira Nunes Pinto, Aline Pereira da Rocha, Keilla Martins Milby, César Ramos Rocha-Filho, Felipe Sebastião de Assis Reis, Nelson Carvas Junior, Vinicius Tassoni Civile, Rodolfo Rodrigo Pereira Santos, Giulia Fernandes Moça Trevisani, Laura Jantsch Ferla, Gabriel Sodré Ramalho, Maria Eduarda Santos Puga, Virgínia Fernandes Moça Trevisani, Alvaro Nagib Atallah

## Abstract

**CONTEXT AND OBJECTIVE:** Coronavirus disease 2019 (COVID-19) has emerged in China in December 2019 and rapidly spread. Although extraordinary efforts have been made on research regarding pharmacological interventions, none have proven effective. This is the protocol for a rapid living systematic review that aims to compare the effectiveness and safety of different pharmacological interventions for the treatment of COVID-19.

**METHODS:** rapid living systematic review methodology with Network Meta-Analysis following the recommendations of Cochrane Handbook. We will include randomized controlled trials (RCT) and quasi-RCTs that evaluate single and/or combined pharmacological interventions at any dose for the treatment of COVID-19. We will search PubMed, Embase, Cochrane Central Register of Controlled Trials (CENTRAL), LILACS, Scopus and SciELO to identify potentially eligible studies. No language restrictions will be used in the selection. We will perform the critical appraisal of included studies with the Risk of Bias tool and the certainty of evidence will be evaluated using the Grading of Recommendations Assessment, Development and Evaluation (GRADE).

## INTRODUCTION

Severe Acute Respiratory Coronavirus 2 (SARS-CoV-2), a novel virus that causes Coronavirus Disease 2019 (COVID-19), has emerged in China in December 2019 and on March 11^th^, 2020, a pandemic was already declared by the World Health Organization (1). As it has recently emerged, much still needs to be understood about this infection. Overall fatality rate, accuracy of diagnostic tests and effectiveness of current treatments are some of the most important and controversial topics.

Fatality rate has varied substantially among countries and possible explanations are the accuracy of the tests and the difference between strategies used for SARS-CoV-2 testing. For example, in countries like Brazil, test availability is currently limited, and tests are prioritized for patients with more severe clinical symptoms who are suspected of having COVID-19 (2). On the other hand, countries such as the Republic of Korea have adopted a strategy of widely testing for SARS-CoV-2. As many patients with mild symptoms (which accounts for nearly 80% of SARS-CoV-2 infections) who would not be tested in Brazil were probably identified and included in the denominator in Korea, a much lower case-fatality rate (1%) compared with Brazil (6.1%) is expected (3), and the actual overall mortality rate may be closer to Korea than Brazil estimations.

Although mortality rate is concerning, the high transmissibility of the disease is much more alarming. Even if a low percentage of patients need hospitalization, the rapid spread of the disease and large number of people infected has overwhelmed the healthcare systems worldwide. To counteract the spread, severe social distancing measures—travel restrictions, closures of schools, and many businesses—are taking an unprecedented socioeconomic and psychological toll. Therefore, COVID-19 has caused an enormous impact on people’s quality of life and posed far-reaching threats, especially to the economy, health, and to the sustainability of healthcare systems (4).

It is, therefore, urgent the need of finding effective interventions to avoid the progression of the disease and unburden the health care systems. Although extraordinary efforts have been made on research regarding pharmacological interventions, none have proven effective. Therefore, we aim to compare the effectiveness and safety of different pharmacological interventions for the treatment of COVID-19. This is the protocol for our rapid living systematic review.

## METHODS

This rapid living systematic review and network meta-analysis (if applicable) protocol was registered in the PROSPERO ‘‘International Prospective Register of Systematic Reviews’’ (CRD42020179818), and was developed following the preferred reporting items for systematic review and meta-analysis protocols (PRISMA-P) guidance (5). The final report will comply with the recommendations of the PRISMA extension statement for reporting of systematic reviews incorporating network meta-analyses (6). To conduct the rapid systematic review, we will employ abbreviated systematic review methods. Compared with the methods of a systematic review, we will not perform independent screens of abstracts and we will not search grey literature (7).

### Design

We will perform a rapid living systematic review methodology with network metaanalysis following the recommendations proposed by the Cochrane Handbook (7). As this will be a living review, it will be continually updated.

### Eligibility Criteria

#### Types of studies

We will include randomized controlled trials (RCT) and quasi-RCTs that evaluate single and/or combined pharmacological interventions at any dose for the treatment of COVID-19.

#### Types of participants

We will include studies with patients with confirmed diagnosis of infection of SARS-CoV-2.

#### Type of interventions

Any pharmacological intervention (used alone or combined with other interventions) for the treatment of COVID-19.

#### Type of comparators

Any other pharmacological interventions, placebo or standard treatment.

#### Outcome measures

- Primary outcomes

- Mortality rate;
- Length of hospital stay;
- Adverse events;
- Secondary outcomes

- Time to clinical improvement;
- Length of Intensive Care Unit stay;
- Number of patients under invasive mechanical ventilation;
- Time to viral clearance;

#### Report characteristics

We will include studies performed since November 2019. No language restrictions will be used in the selection.

### Data Sources and Searches

We will search PubMed, Embase, Cochrane Central Register of Controlled Trials (CENTRAL), LILACS, Scopus and SciELO using relevant descriptors and synonyms, adapting the search to the specifications of each database to identify published, ongoing, and unpublished studies. We will also search the following COVID-19 specific databases: Epistemonikos COVID-19 L·OVE platform; ClinicalTrials.gov; The World Health Organization International Clinical Trials Registry Platform (WHO ICTRP). Finally, we will use the technique of snowballing, searching the lists of references of the included studies. No language restrictions will be used in the selection.

### Search Strategy

We will use the terms related to the problem of interest and the filter for randomized studies provided by Haynes et al. (8). The search strategy in MEDLINE via Pubmed is shown in Table 1.

**Table 1.** Systematic review Search Strategy

| Number | Combiners | Terms |
| --- | --- | --- |
| 1 | Problem of interest | (("Coronavirus"[Mesh]) OR ("Coronaviridae"[Mesh]) OR ("Coronavirus Infections"[Mesh]) OR coronavirinae OR ("COVID-19" [Supplementary Concept]) OR COVID OR ("severe acute respiratory syndrome coronavirus 2"[Supplementary Concept]) OR SARS-CoV-2 OR ("betacoronavirus"[MeSH Terms]) OR Coronaviruses OR 2019-nCoV OR nCoV OR COVID19 OR (Corona virus)) |
| 2 | Type of Study | (clinical[Title/Abstract] AND trial[Title/Abstract]) OR clinical trials as topic[MeSH Terms] OR clinical trial[Publication Type] OR random*[Title/Abstract] OR random allocation[MeSH Terms] OR therapeutic use[MeSH Subheading]) |
| 3 |  | #1 AND #2 AND #3 |
|  | Filters | **Publication date from 2019/11/01** |

The search strategy above will be used in Medline via Pubmed and will be adapted to the specifications of each database.

### Study Selection

Based on pre-specified eligibility criteria, two authors will select the studies for inclusion in the review (ACPNP and APR). When two studies are found in more than one database (duplicated studies) we will consider only one of them for inclusion. If reports using the same participants and different outcome measurements or using different time points for the assessments are found, both reports will be included (the two reports will be considered as parts of only one study). If duplicated reports are found, e.g. studies with the same participants, with the same outcome measurements and using the same time points for the assessments, the report with the smaller sample size will be excluded.

After removing duplicate studies and reports, the authors will read the study titles and abstracts. Studies that clearly do not match the inclusion criteria for the review will be excluded. The selected studies will then be fully read under further scrutiny; the reasons for their exclusion will be presented. Disagreements between authors regarding study inclusion will be resolved by the third author (ANA). To optimize the process of screening and selection of studies, we will use Rayyan application (9).

### Data Extraction

Two authors (ACPNP and APR) will independently extract data. Discrepancies or disagreements will be solved by a third author (CRRF). We will use a predefined form to extract data from included studies. The form will have information related to: - the patients (demographic and clinical characteristics); - the pharmacological treatment (name of the drug, treatment duration; dose); - time points used for the assessments; - number of patients lost to follow-up (in each group); - reasons for loss to follow-up; - approach for handling missing data (data imputation/how data imputation was performed, use of intention-to-treat approach); - sources of funding; - possibility of conflict of interests; - adverse events; - outcome measures; - protocol deviations.

To assess the feasibility of performing a meta-analysis, we will also extract the following data for each primary and secondary outcome measure: - total number of patients (in each group); - number of events in each group (for dichotomous outcomes); - mean, standard deviation, standard error, median, interquartile range, minimum, maximum, 95% confidence interval (CI) (for continuous outcomes); - p-value.

### Assessment of Methodological Quality in included studies and certainty of evidence

We will perform critical appraisal of included studies with Risk of Bias tool (10) as recommended by Cochrane Collaboration. We will evaluate the certainty of evidence using the Grading of Recommendations Assessment, Development and Evaluation (GRADE) (11). GRADE judgement is based on the overall risk of bias, consistency of the results, directness of the evidence, publication bias and precision of the results for each outcome. The GRADE profiler software, available online, will be used to summarize our findings on the certainty of evidence (12). Assessment of risk of bias (ACPNP and KMMM), and assessment of the certainty of evidence (VTC and NCJ) will be performed by two review authors. All the disagreements in the assessment of the risk of bias or the certainty of evidence will be solved through discussion or, if required, by consulting with a third author (ANA).

### Data analysis

#### Intervention network geometry

We will use the forest.netmeta function of the netmeta package to build and present the geometry of different interventions. We will use Nodes to represent the intervention and edges to show comparisons between interventions.

#### Network meta-analysis

We plan to use the netmeta package version 1.2-1 implemented in R-3.6.2 software for Mac to perform a network meta-analysis and synthesize the direct and indirect evidence of the therapeutic effect of the pharmacological therapeutic effects. We will use the node splitting method to assess an inconsistency between direct and indirect comparisons when there is a loop connecting three arms. We will present the treatment ranking by *P*-scores based on the points estimates and standard error of the available network.

#### Subgroup and sensitivity analysis

In case of possible significant heterogeneity or inconsistency, we will use subgroup analysis. We will explore, when possible, the following available variables: age, sex, comorbidities, strategy to address pandemic threat (e.g. quarantine, lockdown, social distance) and disease severity. We will also perform sensitivity analysis for include studies with high risk of bias, missing data.

#### Publication bias

To investigate the influence of small-studies effects, we will use the visual inspection method of funnel plot when at least ten studies were included in a meta-analysis, followed by Egger’s test (10).

## DISCUSSION

This rapid living review will systematically evaluate the best available evidence on the pharmacological treatment of COVID-19, which we expect will help the front line on their decision-making processes. Currently, there is no evidence from RCT that has shown potential improvements in outcomes in patients with confirmed COVID-19. In addition to the fact that there is no drug with proven efficacy or approval by drug regulatory agencies for the treatment of COVID-19. However, to date, there are almost two thousand ongoing trials on SARS-CoV-2 infection registered in WHO ICTRP. While different therapeutic classes emerge as possible treatments against COVID-19, the availability of drug-related information, time to clinical improvement, adverse events and mortality rate are still challenging. There is an urgent need to find reliable evidence on which pharmacological treatment is more effective and safer for treating SARS-CoV-2 infection.

To ensure the quality of the results, we will follow the Cochrane Handbook of Systematic Reviews recommendations (10). We believe this rapid systematic review with network meta-analysis and extensive searches will be able to summarize the current available evidence on pharmacological treatments and to provide important information for clinical decision-making on COVID-19 that has recently emerged and caused a deadly pandemic.

## Data Availability

The authors confirm that the data supporting the findings of this study are available within the article

## REFERENCES

1. World Health Organization. Coronavirus disease (COVID-19) outbreak situation 2020. Available from: https://www.who.int/emergencies/diseases/novel-coronavirus-2019

2. Brazilian Health Ministry. Infecção Humana pelo Novo Coronavírus (2019-nCoV). Boletim Epidemiologico 2. 2020.

3. Rajgor DD, Lee MH, Archuleta S, Bagdasarian N, Quek SC. The many estimates of the COVID-19 case fatality rate. Lancet Infect Dis 2020.

4. World Health Organization. Coronavirus Disease (COVID-19) Pandemic. Available online: https://www.who.int/emergencies/diseases/novel-coronavirus-2019 (accessed on April 29th.

5. Moher D, Shamseer L, Clarke M, Ghersi D, Liberati A, Petticrew M, et al. Preferred reporting items for systematic review and meta-analysis protocols (PRISMA-P) 2015 statement. Syst Rev. 2015;4:1. DOI: 10.1186/2046-4053-4-1.

6. Hutton B, Salanti G, Caldwell DM, Chaimani A, Schmid CH, Cameron C, et al. The PRISMA extension statement for reporting of systematic reviews incorporating network meta-analyses of health care interventions: checklist and explanations. Ann Intern Med. 2015; 162(11):777–84

7. Garritty C, Gartlehner G, Kamel C, King VJ, Nussbaumer-Streit B, Stevens A, et al. Cochrane Rapid Reviews Interim Guidance from the Cochrane Rapid Reviews Methods Group 2020. Available from: https://methods.cochrane.org/rapidreviews/sites/methods.cochrane.org.rapidreviews/files/public/uploads/cochrane_rr_-_guidance-23mar2020-v1.pdf.

8. Haynes RB, McKibbon KA, Wilczynski NL, Walter SD, Werre SR. Optimal search strategies for retrieving scientifically strong studies of treatment from Medline: analytical survey. BMJ. 2005

9. Ouzzani M, Hammady H, Fedorowicz Z, Elmagarmid A. Rayyan—a web and mobile app for systematic review. Systematic Reviews. 2016; 5: 210.

10. Higgins J. Cochrane Handbook for Systematic Reviews of Interventions 2011.

11. Atkins D, Best D, Briss PA, Eccles M, Falck-Ytter Y, Flottorp S, et al. Grading quality of evidence and strength of recommendations. Bmj. 2004;328(7454):1490. DOI: 10.1136/bmj.328.7454.1490.

12. GDT G. Grade’s software for summary of findings tables, health technology assessment and guidelines 2015 [Available from: https://gradepro.org/.

